# Optimizing Hepatitis C Virus Testing in the Era of Point-of-Care RNA Diagnostics

**DOI:** 10.1101/2025.08.25.25334368

**Authors:** Emily W. Helm, H. Nina Kim, Alexander L. Greninger

## Abstract

Major advances in hepatitis C virus (HCV) treatment have made timely and accurate diagnosis a critical determinant for United States elimination goals. Recently authorized point-of-care (POC) HCV RNA testing enables faster diagnosis than the traditional two-step algorithm but comes with higher laboratory costs. We analyzed HCV testing data from 2017-2024 across three medical centers in Seattle, Washington, to evaluate strategies for integrating new rapid direct detection tests. HCV antibody testing volumes increased 72% over the study period, with outpatient settings accounting for 76.0% of tests and a 2.7% positivity rate. Emergency department testing increased by 682% to 5,654 tests in 2024, with a 10.3% positivity rate, such that one-third of all HCV diagnoses in our medical system now originate from the public county hospital emergency department. Adoption of sample-to-answer HCV RNA testing in 2024 reduced median collection-to-result turnaround times for antibody-positive specimens from 84 hours (IQR 58 - 120) to 45 hours (IQR 28 - 57). Using a viral load cut-off of 10,000 IU/mL, HCV antigen testing was estimated to detect 98% of infections. Converting all HCV testing to POC RNA would increase laboratory costs by 260% (+$6,439 per HCV infection detected), while restricting POC RNA to the public county hospital emergency department would increase costs by 22.3% (+$552 per HCV infection detected). Reflexing antibody-positive samples to antigen testing slightly reduced costs. These findings highlight the significant laboratory costs associated with POC HCV RNA testing and the need for specific reimbursement and funding mechanisms for new HCV testing algorithms.

**Importance:** Hepatitis C virus (HCV) elimination in the United States requires rapid and reliable diagnosis, yet current testing pathways are too slow for treatment initiation in a single clinical visit. Analysis of more than 325,000 HCV test results from 2017-2024 from our academic medical system in Seattle, Washington highlights the growing role of emergency departments, particularly those serving safety-net populations, in making new HCV diagnoses. While point-of-care HCV RNA testing can enable connection to treatment, it substantially increases laboratory costs when implemented broadly. Targeted use of point-of-care HCV RNA in high-yield settings such safety-net emergency departments is essential to maximize public health impact while preserving laboratory resources. These findings highlight the need for policy and reimbursement frameworks that support cost-effective deployment of new HCV diagnostic technologies.

## Introduction

Hepatitis C virus (HCV) remains a major public health concern, with an estimated prevalence of 1.6% among adults in the United States^1^. With the opioid epidemic, rates of acute hepatitis C cases in the United States tripled from 2009 to 2018^2^ as injection drug use became the most identified risk factor for HCV infection^3^. The treatment landscape for HCV has undergone a dramatic transformation over the past decade. The introduction of direct-acting antivirals has revolutionized care, offering short, all-oral regimens that cure over 90% of patients regardless of genotype or liver disease stage ^4–6^. Despite the availability of curative therapy, the CDC estimates that 69,000 acute HCV infections occurred in 2023, along with 101,525 newly reported chronic HCV cases^7^.

The pace of HCV elimination now hinges on improvements in case-finding and diagnostics that link patients to care. Recent modeling estimates that only 60% of people infected with HCV in the United States are aware of their status^2^. Among those who received HCV testing in the United States between 2013-2021, only 34% had cleared infection, indicating that many patients are not being successfully connected to treatment^8^. These data underscore the urgent need for an effective testing paradigm that identifies active infections and facilitates connection of patients to treatment.

The current recommended HCV screening algorithm begins with antibody testing, followed by reflex HCV RNA testing if the antibody result is reactive^9^. Because antibody testing costs roughly one-third that of RNA testing based on current Centers for Medicare & Medicaid Services (CMS) fee schedules^10^, this approach offers a cost-effective method for broad population screening but results in delayed detection as well as operational complexity. Positive antibody results require confirmation with RNA testing before treatment, as approximately 37% of infected individuals may clear the virus spontaneously^11^, and more than 1.8 million Americans have been treated for HCV and remain antibody-positive^12^. Positive antibody tests can also be confirmed using the slightly less sensitive HCV core antigen test ^13,14^. In 2023, the World Health Organization endorsed either assay for confirming a positive antibody screen^15^; however, no HCV antigen test has yet received FDA authorization^16^.

While HCV screening ideally occurs in primary care settings where appropriate follow-up can be arranged, many individuals lack access to routine primary care and instead rely on emergency departments for care^17^. Factors associated with not having a primary care physician – including younger age, non-private insurance status, experiencing homelessness, and having a substance use disorder – often overlap with risk factors for HCV infection^18,19^. The traditional two-step algorithm of HCV testing generally does not allow confirmatory testing to be completed during a single emergency department encounter. Recently, the FDA approved the first point-of-care (POC) HCV RNA test for HCV infection screening^20^, enabling detection of active HCV infection and potential initiation of therapy during the emergency department visit. A recent large randomized controlled trial of HCV antibody screening in emergency departments found that only 16% of patients with new HCV diagnoses initiated antiviral therapy when only antibody results were available during the emergency department encounter, underscoring the importance of having RNA results available in real time^21^. However, POC HCV RNA testing is limited by cost and testing capacity^22^. Determining how to implement POC HCV RNA testing effectively is an important question for medical systems and clinical microbiology labs.

Here, we analyzed HCV testing from 2017-2024 across our medical system to inform the implementation of HCV RNA screening. Our academic medical system includes three hospitals with distinct patient populations as well as outpatient services and a cancer center, thus allowing us to analyze testing patterns across different settings. In addition, we performed cost modeling to more accurately estimate overall laboratory costs associated with different testing approaches.

## Materials and Methods

### Clinical and testing setting

Located within King County, Washington, University of Washington (UW) Medicine comprises three hospitals, a cancer center, and a network of clinics. These hospitals include a 660-bed academic medical center hospital, a 500-bed public county hospital, and a 281-bed community hospital, each with its own emergency department. UW Medicine operates 24 primary care clinics and multiple outpatient sites, reporting 2,231,037 clinic visits in 2024^23^. Overall, King County reported 1,254 newly diagnosed cases of HCV infection in 2019 and 755 in 2024 ^24^. In 2019, the highest rates of HCV diagnoses per 100,000 population were among individuals aged 60–69, followed by those aged 30–39, 50–59, and 40–49. By 2024, the age groups with the highest rates remained 60–69, followed by 40–49, 30–39, and 50–59^24^.

### UW Medicine HCV testing

Both HCV antibody and viral load tests are offered through the UW Virology laboratory at a location that is at least one mile off-site from UW Medicine hospitals and clinics. A courier system is in place to bring patient samples from other sites to the laboratory. HCV testing can be ordered as standalone antibody or viral load testing, or as reflex viral load testing following a reactive antibody test. Antibody tests are performed using the Abbott Architect i2000. Viral load testing was performed on the Abbott m2000 from 2017-2024, switching to the Hologic Panther on November 1, 2024.

### HCV testing data

HCV testing data from January 1, 2017 to December 31, 2024, were obtained through the UW Medicine Department of Laboratory Medicine and Pathology Data Warehouse. Hepatitis C antibody and RNA testing data for both reflexed and unreflexed testing were identified using test order codes. The dataset was filtered to exclude proficiency testing, research testing, and non-patient samples. Orders without results were also excluded to avoid analyzing cancelled or reordered samples. For analysis of ordering locations, location codes were grouped into seven categories: employee health, cancer center, outside (reference laboratory services for external non-UW clients), emergency department (ED), inpatient, outpatient, and other. The “other” category primarily includes infusion clinics and outpatient procedure locations that did not fit into other groups. All analyses were performed using R version 4.5.1. This study was approved by the University of Washington Institutional Review Board with a consent waiver (STUDY00010205).

### Cost Projections

We used two approaches to estimate laboratory costs associated with HCV testing. The first approach utilized reimbursements from the 2025 CMS Clinical Laboratory Fee Schedule: $14.27 per HCV antibody test and $42.84 per HCV RNA test^10^. The second approach relied on internal cost accounting data from the UW Department Laboratory Medicine and Pathology of $23.09 per HCV antibody test and $49.16 per HCV RNA viral load test. For HCV POC RNA testing, the cost was estimated at $90.69 per test, based on reagent costs ($50) combined with internal staffing costs for nucleic acid amplification tests (NAAT) performed one specimen at a time. Potential HCV antigen test costs were approximated using reagent costs for qualitative hepatitis B surface antigen testing ($4.64 per test). For combined antigen/antibody testing, this value was added to the antibody test cost of $27.73 per test, assuming no additional staffing time. For antibody testing reflexed to antigen testing, an additional $5.03 per test was added to antibody test cost to cover reagents and the minimal extra staff time for processing the reflex test.

Projected testing volumes for different algorithms were based on HCV testing volumes and positivity rates observed at UW Medicine in 2024. To accurately capture orders used for HCV screening, only tests ordered as “HCV antibody with reflex to PCR” were included. Cost differences between algorithms were calculated by comparing the total costs of each scenario to the cost of maintaining the traditional two-step algorithm. To simplify modeling of the sensitivity of the HCV antigen test, we assumed 100% sensitivity at viral loads >10,000 IU/mL and no sensitivity at viral loads < 10,000 IU/mL^13,14^.

## Results

### Hepatitis C Antibody Utilization

As the CDC currently recommends a two-step algorithm for HCV screening, we first analyzed ordering trends for HCV antibody testing across our academic medical system. HCV antibody testing volumes increased from 26,188 tests in 2017 to 45,010 tests in 2024, including a 1.9-fold increase between 2019 and 2021 corresponding both to a UW initiative to increase ED-based HCV testing as well as updated 2020 CDC guidance recommending one-time HCV screening for all adults^25^ (Figure 1A). With the increased testing, the positivity rate declined from 7.2% in 2019 to 4.2% in 2024 (Figure 1B). The HCV antibody testing population became noticeably younger between 2017 and 2024, the proportion of individuals age 20-39 years almost doubling from 26.9% to 51.8% (Table 1). Between 2017 and 2024, outpatient settings accounted for 76.0% of antibody tests, followed by inpatient (8.5%) and the emergency department (6.9%) settings (Figure 1C). Notably, emergency department testing rose 7.8-fold from 723 tests in 2017 to 5,654 tests in 2024 (Figure 1C). Positivity rates were lowest in outpatient testing (2.7%) and highest in inpatient (17.0%) and emergency department (13.7%) testing (Figure 1D).

**Figure 1.**
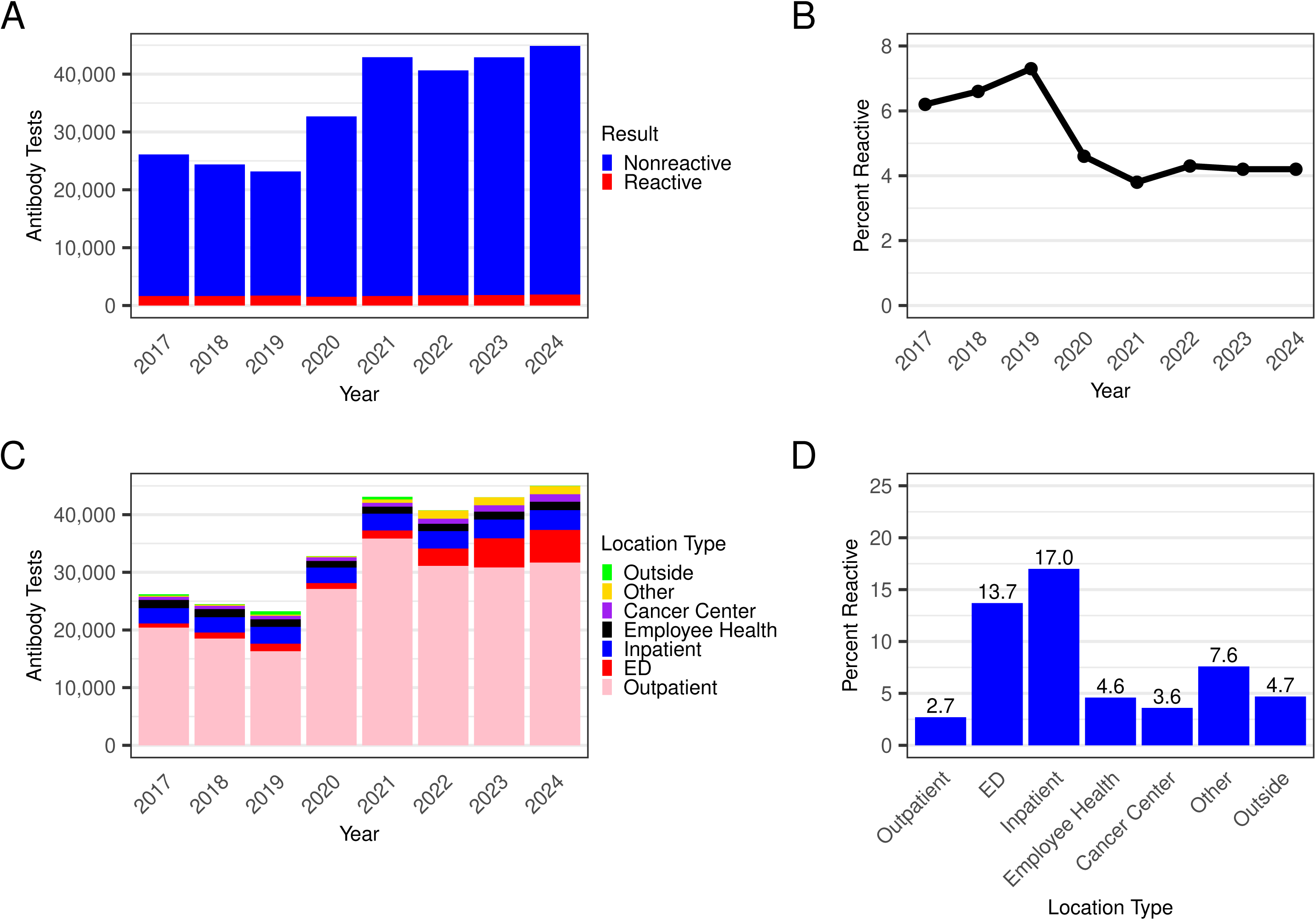
HCV antibody tests are primarily ordered from outpatient locations and have increased 1.7-fold from 2017 to 2024. (A) Number of HCV antibody tests per year across all sites summarized by test result. Indeterminate results are not shown. (B) Percent positivity rate of HCV antibody tests per year across all sites. (C) Number of HCV antibody tests per year by location. (D) Percent of reactive HCV antibody tests from 2017 to 2024 by location.

**Table 1.** Demographic Information for HCV Antibody Testing by Year.

|  | 2017 | 2018 | 2019 | 2020 | 2021 | 2022 | 2023 | 2024 |
| --- | --- | --- | --- | --- | --- | --- | --- | --- |
| **Age Group** |  |  |  |  |  |  |  |  |
| <10 | 0.5 | 0.3 | 0.3 | 0.1 | <0.1 | 0.1 | 0.1 | 0.1 |
| 10-19 | 1.2 | 1.3 | 1.5 | 1.3 | 1.4 | 1.6 | 1.9 | 2.0 |
| 20-29 | 11.6 | 12.9 | 13.9 | 18.4 | 21.2 | 22.8 | 22.3 | 21.7 |
| 30-39 | 15.3 | 17.2 | 19.9 | 25.1 | 28.5 | 29.2 | 29.4 | 30.1 |
| 40-49 | 11.1 | 11.8 | 13.2 | 18.2 | 17.9 | 16.9 | 16.9 | 16.8 |
| 50-59 | 26.3 | 23.9 | 20.6 | 16.9 | 15.4 | 13.6 | 13.2 | 12.6 |
| 60-69 | 26.1 | 24.1 | 21.3 | 11.4 | 9.2 | 9.9 | 9.9 | 10.2 |
| 70-79 | 6.8 | 7.3 | 7.8 | 7.7 | 5.6 | 5.0 | 5.2 | 5.0 |
| 80-89 | 1.1 | 1.0 | 1.3 | 0.7 | 0.7 | 0.9 | 1.0 | 1.2 |
| ≥90 | 0.1 | 0.1 | 0.1 | 0.1 | 0.1 | 0.1 | 0.1 | 0.2 |
| Unknown | 0.1 | 0.1 | 0.1 | 0.1 | <0.1 | 0.1 | 0.1 | 0.1 |
| **Race** |  |  |  |  |  |  |  |  |
| White | 44.3 | 42.4 | 47.4 | 55.2 | 59.9 | 60.1 | 58.3 | 56.8 |
| Unknown | 41.6 | 41.7 | 31.0 | 15.5 | 5.6 | 4.1 | 5.7 | 7.2 |
| Asian | 6.7 | 7.4 | 8.2 | 13.8 | 17.5 | 17.7 | 17.5 | 17.3 |
| Black or African American | 3.6 | 3.7 | 7.1 | 7.6 | 8.5 | 9.5 | 10.8 | 11.5 |
| Other Race | 2.7 | 3.4 | 4.2 | 5.6 | 6.1 | 6.1 | 5.2 | 4.5 |
| American Indian or Alaska Native | 0.8 | 0.9 | 1.4 | 1.4 | 1.5 | 1.6 | 1.6 | 1.7 |
| Native Hawaiian or Other Pacific Islander | 0.4 | 0.6 | 0.7 | 0.7 | 0.8 | 0.9 | 0.9 | 0.9 |
| **Sex** |  |  |  |  |  |  |  |  |
| F | 49.9 | 48.3 | 48.4 | 52.7 | 54.0 | 53.1 | 52.9 | 52.4 |
| M | 49.8 | 50.8 | 50.8 | 46.7 | 45.4 | 46.2 | 46.4 | 46.4 |
| Unknown/Other | 0.3 | 0.9 | 0.8 | 0.6 | 0.6 | 0.7 | 0.8 | 1.1 |
| **Patient Zip Code** |  |  |  |  |  |  |  |  |
| 981 | 38.3 | 38.3 | 38.7 | 44.8 | 50.3 | 51.6 | 52.6 | 53.1 |
| 980 | 27.1 | 25.6 | 23.1 | 26.3 | 26.3 | 25.2 | 25.2 | 25.3 |
| 982 | 9.4 | 10.5 | 10.4 | 8.4 | 6.6 | 6.4 | 6.2 | 6.0 |
| Unknown | 6.5 | 6.1 | 8.2 | 3.9 | 3.2 | 3.2 | 3.2 | 3.4 |
| 983 | 4.3 | 4.9 | 4.4 | 3.6 | 3.4 | 3.4 | 3.5 | 3.6 |
| 985 | 3.4 | 3.6 | 3.5 | 3.8 | 1.6 | 1.7 | 1.7 | 1.6 |
| 984 | 2.2 | 2.4 | 2.4 | 2.3 | 2.0 | 2.2 | 2.1 | 2.0 |
| 989 | 1.1 | 1.0 | 1.2 | 0.8 | 0.8 | 0.9 | 0.9 | 0.8 |
| 988 | 0.7 | 0.7 | 0.7 | 0.5 | 0.5 | 0.4 | 0.4 | 0.4 |
| 993 | 0.6 | 0.6 | 0.6 | 0.5 | 0.5 | 0.4 | 0.5 | 0.5 |
| All Others | 6.4 | 6.3 | 6.8 | 5.1 | 4.8 | 4.6 | 3.7 | 3.3 |

Given the increase in HCV antibody testing from the emergency department, we further characterized testing at this location. Between 2017 and 2024, the public county hospital emergency department accounted for 73.3% of all ED HCV antibody tests, followed by the academic medical center (18.7%) and community hospital (8.0%) (Figure 2A). Since the public county hospital contributed nearly three-quarters of ED tests, we focused subsequent analyses on this site. From 2021 to 2023, the public county hospital emergency department rose 3.7-fold, from 1004 tests in 2021 to 3,751 tests in 2023, reaching 4,013 tests in 2024 (Figure 2B). During this time period, the positivity rate declined from 17.0% in 2021 to 12.5% in 2023 (Figure 2C).

**Figure 2.**
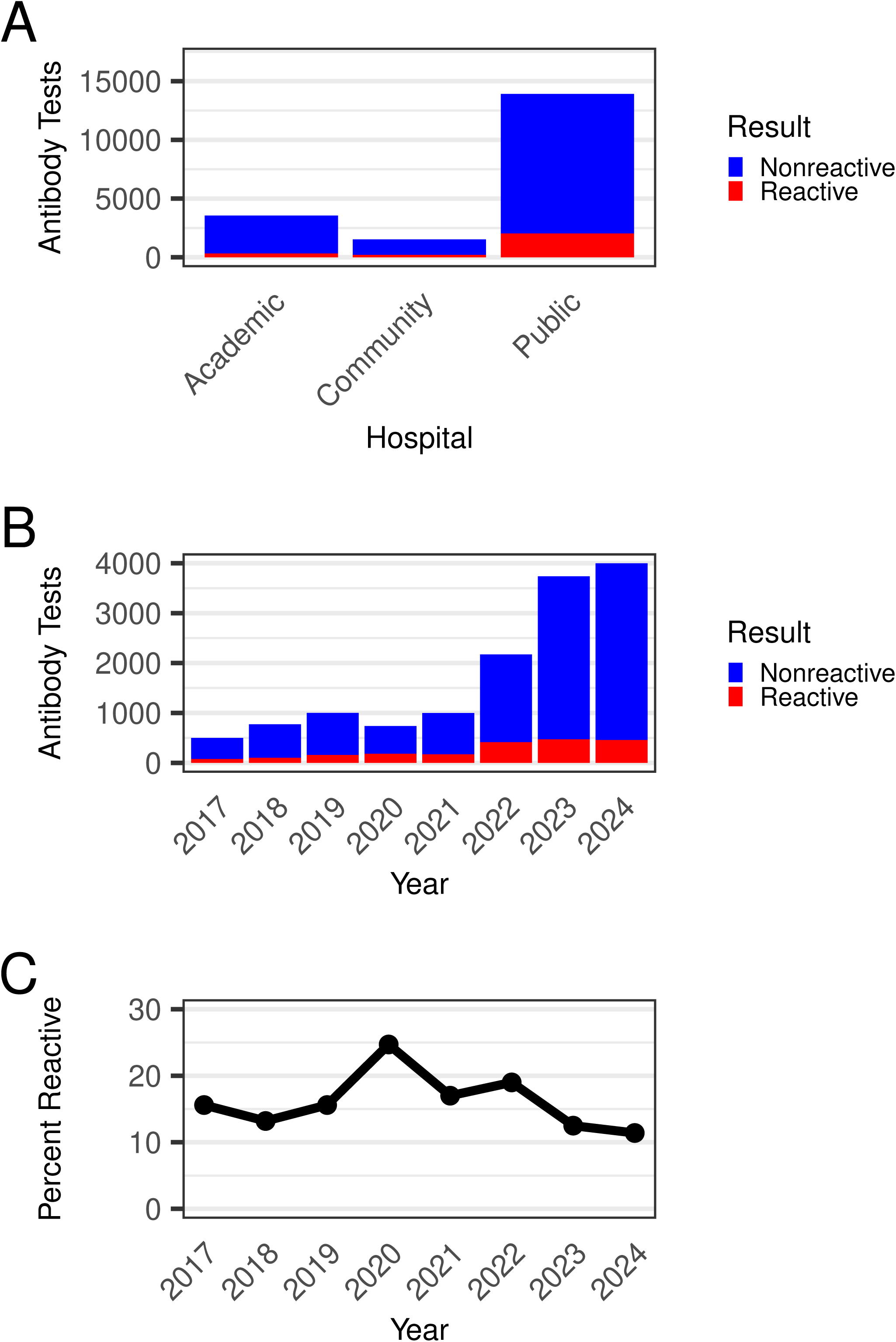
HCV antibody tests from the emergency department are overwhelmingly from the public county hospital emergency department and have increased 8.0-fold from 2017 to 2024. (A) Number of HCV antibody tests originating from the emergency department from 2017-2024 summarized by test result and hospital site. Indeterminate results were not included in the analysis. (B) HCV antibody tests originating from the public county hospital emergency department were analyzed by year and result. Indeterminate results were not included in the analysis. (C) Percent of reactive HCV antibody tests originating from the public county hospital emergency department per year.

### HCV Viral Load Utilization

HCV antibody testing is only the first step in the CDC algorithm, as all reactive antibody tests should be confirmed with viral load testing to quantify HCV RNA levels^25^. Unlike antibody testing, HCV RNA tests decreased by 35.3% from 2017 to 2024, with steady declines during the 2020 pandemic year followed by stabilization from 2021 to 2024 (Figure 3A). In 2017, outpatient settings accounted for 64.4% of viral load tests, decreasing to 37.3% in 2024 (Figure 3A). Alongside the overall decrease in viral load testing, the proportion of tests with detectable HCV RNA fell from 38.5% in 2017 to 15.1% in 2024 (Figure 3B). Conversely, inpatient and emergency department viral load testing increased from 19.4% of all HCV RNA tests in 2020 to 38.6% by 2024 (Figure 3A). Viral load testing in the public county hospital emergency department rose 6.6-fold between 2017 and 2024 (Figure 3C). Despite the increased volume, the positivity rate for detectable viral loads from the public county hospital emergency department remained high at 30.7% in 2024 (Figure 3C).

**Figure 3.**
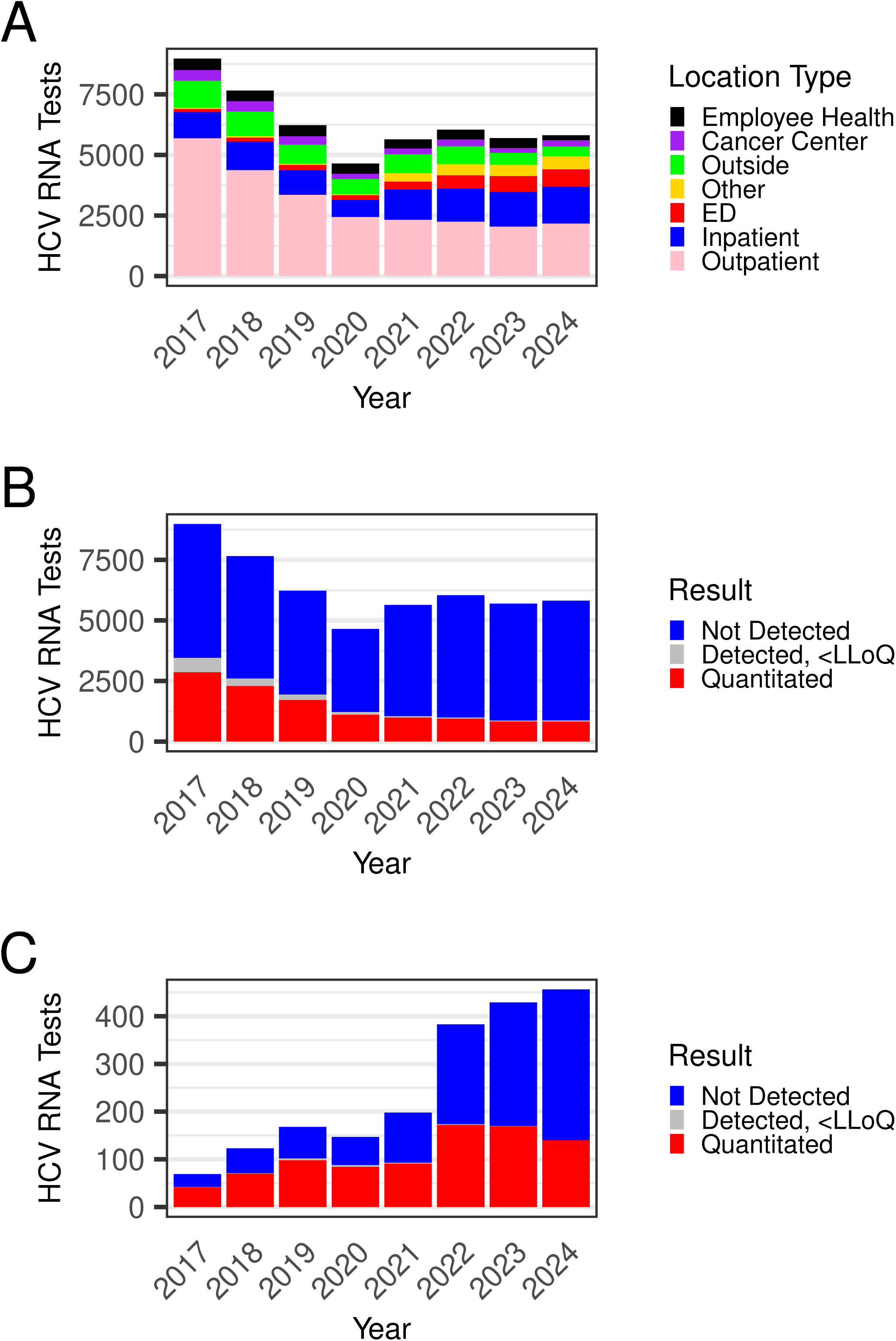
The number of HCV RNA tests decreased overall but increased in the public county hospital emergency department from 2017 to 2024. (A) Number of HCV RNA tests per year by location type. (B) Number of HCV RNA tests across all locations per year summarized by test result. (C) Number of HCV RNA tests from the emergency department of the public county hospital per year summarized by test result.

### Turnaround Time Analysis

We next examined turnaround times for reflex HCV antibody and viral load tests. From January 2017 to October 2024, HCV viral load testing was performed twice a week using the multi-step Abbott m2000 platform, resulting in a median collection-to-RNA-result turnaround time of 84 hours (IQR 58-120) for reactive screening tests (Figure 4). In November 2024, viral load testing switched to the Hologic Panther platform, enabling three runs per week. However, true random-access testing remained limited due to quality control costs associated with quantitative viral load testing. This change reduced the median turnaround time to 45 hours (IQR 28-57) (Figure 4).

**Figure 4.**
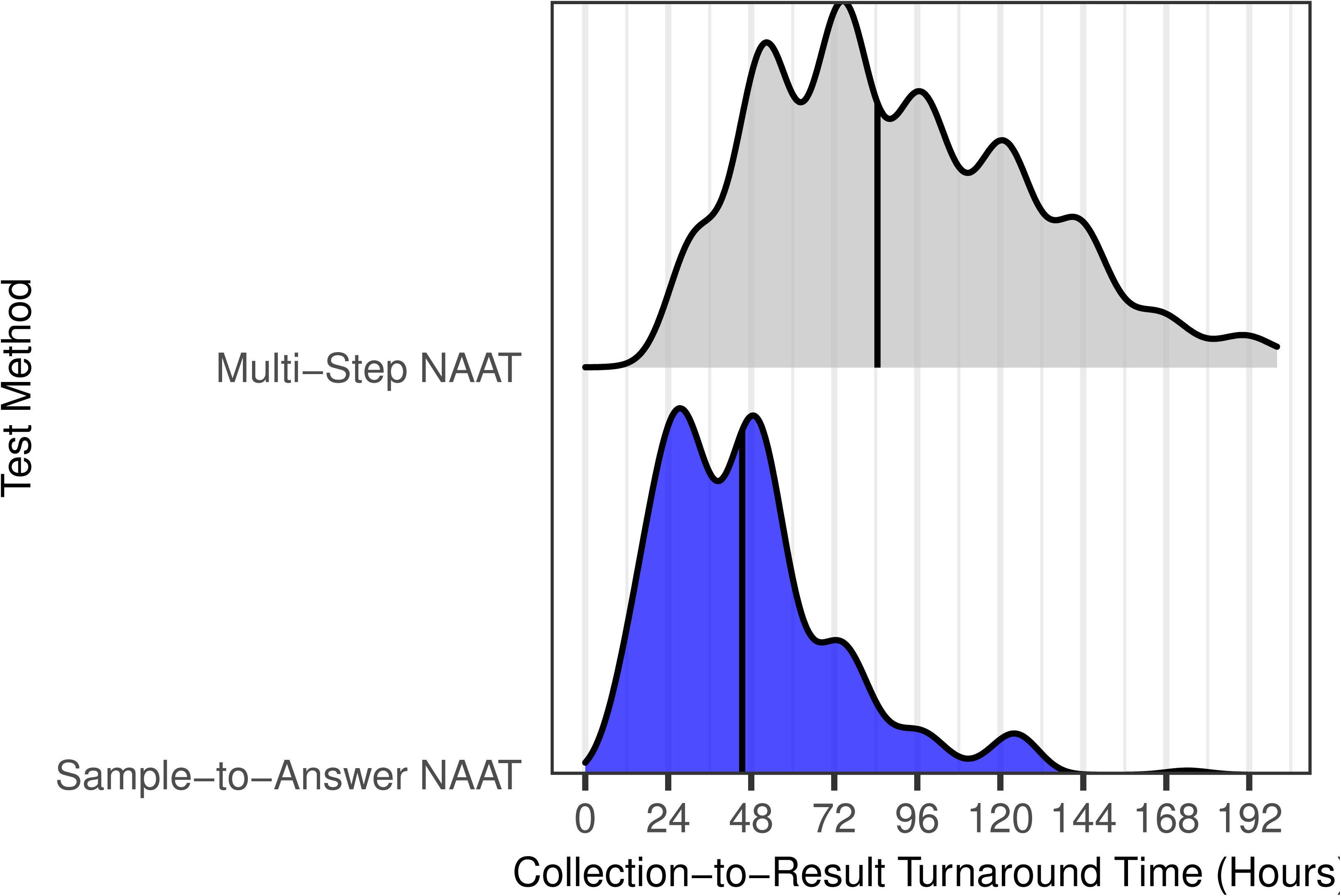
Converting HCV viral load testing to random access, sample-to-answer instrumentation has reduced turnaround time from 84 to 45 hours for reflex testing. Turnaround time was analyzed for tests consisting of a positive HCV antibody screen that reflexed to an HCV viral load test. Turnaround time was defined as time from initial collection to HCV viral load result. A ridgeplot is shown comparing the turnaround time while the lab was utilizing batched testing for HCV viral load testing (Multi-Step NAAT) compared to sample-to-answer testing NAAT for HCV viral load. The vertical line represents the median value.

### Cost Projections

Based on this testing data, we examined five scenarios for implementing POC HCV RNA testing instead of initial antibody testing, using 2024 data: 1) all HCV testing, 2) inpatient and emergency department testing only, 3) emergency department testing only, 4) public county hospital emergency department testing only, and 5) maintaining all testing remaining as the traditional two-step algorithm. To simplify projections, we assumed all algorithms would eventually identify all 446 HCV infections detected in 2024. To estimate costs, we first used the 2025 Medicare reimbursement rates for HCV antibody testing and HCV viral load quantification (Table 2). If no changes were made and all testing remained as a two-step algorithm, the total laboratory cost was $704,848, with a laboratory cost per HCV infection detected of $1,580. Switching to single-step POC HCV RNA testing for all screening would increase costs by an additional $1,172,744. Restricting single-step POC HCV RNA testing to inpatient and emergency department settings would increase costs by $211,703; to emergency departments at all three hospitals by $135,450; and to the public county hospital emergency department by $94,051. Corresponding costs per HCV infection detected would be $4,210 for universal single-step RNA testing; $2,055 for inpatient and emergency department testing; $1,884 for all emergency department testing; and $1,791 for the public county hospital emergency department.

**Table 2.** Cost analysis for switching to one-step HCV viral load testing at different implementation locations. The cost difference between using a two-step algorithm for HCV screening compared to single-step HCV RNA testing was compared. The first value listed is based on current Medicare reimbursement rates for HCV antibody and viral load testing and the second value listed is based on estimated lab costs for HCV antibody testing, traditional HCV RNA testing, and point-of-care HCV RNA testing. All values are in US dollars. All values are based on 2024 data for reflex test orders, and costs are estimated for one year.

| Testing converted to single-step HCV RNA | No additional testing | Public County Hospital ED | ED | Inpatient and ED | All Testing |
| --- | --- | --- | --- | --- | --- |
| **Number of one-step HCV RNA tests** |  |  |  |  |  |
|  | 0 | 3,970 | 5,600 | 8,967 | 43,828 |
| **Remaining two-step algorithm tests** |  |  |  |  |  |
|  | 43,828 | 39,858 | 38,228 | 34,861 | 0 |
| **RNA positivity (%) in one-step HCV RNA testing** |  |  |  |  |  |
|  | N/A | 3.75 | 3.40 | 3.88 | 1.02 |
| **Antibody positivity (%) in population undergoing two-step testing** |  |  |  |  |  |
|  | 4.23 | 3.51 | 3.35 | 2.34 | N/A |
| **Laboratory cost of one-step HCV RNA tests** |  |  |  |  |  |
| Medicare | 0 | \$170,075 | \$239,904 | \$384,146 | \$1,877,592 |
| Cost accounting | 0 | \$360,039 | \$507,864 | \$813,217 | \$3,974,761 |
| **Laboratory cost of two-step algorithm testing** |  |  |  |  |  |
| Medicare | \$704,848 | \$628,824 | \$600,394 | \$532,404 | 0 |
| Cost accounting | \$1,103,127 | \$989,231 | \$945,661 | \$845,033 | 0 |
| **Total laboratory cost** |  |  |  |  |  |
| Medicare | \$704,848 | \$798,899 | \$840,298 | \$916,551 | \$1,877,592 |
| Cost accounting | \$1,103,127 | \$1,349,270 | \$1,453,525 | \$1,658,250 | \$3,974,761 |
| **Increased laboratory costs compared to two-step algorithm** |  |  |  |  |  |
| Medicare | \$0 | \$94,051 | \$135,450 | \$211,703 | \$1,172,744 |
| Cost accounting | \$0 | \$246,143 | \$350,397 | \$555,123 | \$2,871,634 |
| **Laboratory cost per detected infection** |  |  |  |  |  |
| Medicare | \$1,580 | \$1,791 | \$1,884 | \$2,055 | \$4,210 |
| Cost accounting | \$2,473 | \$3,025 | \$3,259 | \$3,718 | \$8,912 |

While these calculations provide a framework for reimbursement costs, they do not fully capture laboratory expenses, as currently authorized POC HCV RNA testing is more costly than traditional RNA methods due to higher reagent prices and increased staffing needs. Additionally, reimbursement rates do not always reflect the actual costs of performing tests. Therefore, we conducted a second round of laboratory cost projections using internal UW cost accounting data for HCV antibody and traditional RNA testing, combined with estimated reagent and staffing costs for POC RNA testing (Table 2). Using these internal estimates, the total laboratory cost for HCV testing in 2024 was $1,103,127. We projected added costs of $2,871,634 if all screening shifted to POC HCV RNA testing, compared to $555,123 for inpatient and emergency department testing, $350,397 for all emergency department testing, and $246,143 for testing restricted to the public county hospital emergency department.

While HCV RNA testing is currently the only FDA-authorized method to confirm active HCV infection in the United States, HCV core antigen testing is used in other countries for this purpose^15,26–28^. We therefore projected the costs of implementing either combination HCV antigen/antibody or reflex HCV antibody-to-antigen testing, which could offer lower-cost alternatives if FDA-authorized tests become available in the United States. Because sensitivity is a major concern with antigen testing, we first assessed how many infections would be detectable based on literature indicating antigen tests reliably detect viral loads above 10,000 IU/mL^13,14^. Restricting analysis to viral loads ordered reflexively after positive antibody screening in 2024, 98.1% of detectable HCV RNA tests had viral loads above this threshold and would likely be identified by antigen testing (Figure S1). For combination antigen/antibody testing, we assumed the test would be performed upfront, with any positive antibody and negative antigen results reflexed to RNA testing to ensure infection detection^27^. Using Hepatitis B surface antigen reagent costs as a proxy for HCV antigen costs, we estimated that initiating all screening with combination HCV antigen/antibody tests would add $183,193 in laboratory costs, compared to $25,573 if restricted to inpatient and emergency department testing, $17,169 if restricted to all emergency department testing, and $11,494 if restricted to public county hospital emergency department testing (Table 3). For reflex antibody-to-antigen testing, we assumed that any specimen with an antibody-positive, antigen-negative result would be reflexed to an HCV viral load test to not miss infections. Based on this scenario, we estimated a cost savings of $10,844 if all positive screening antibody tests were reflexed to antigen testing versus a savings of $10,811 for inpatient and emergency department testing, $5,933 for all emergency department testing, and $4,652 for the public county hospital emergency department testing only (Table 4).

**Table 3.** Cost analysis for switching to combination HCV antigen/antibody testing based on implementation locations. The cost difference between using a two-step algorithm for HCV screening compared to initial testing with a combination HCV antigen/antibody test was compared. All values are based on estimated lab costs for HCV antibody testing, traditional HCV RNA testing, and Hepatitis B surface antigen testing which was utilized to estimate HCV antigen costs. All costs are in US dollars. All values are based on 2024 volumes for reflex test orders, and costs are estimated for one year.

| Testing converted to<br>combo antibody/antigen | No additional<br>testing | Public<br>County<br>Hospital ED | ED | Inpatient and<br>ED | All Testing |
| --- | --- | --- | --- | --- | --- |
| **Number of combination antigen/antibody tests** |  |  |  |  |  |
|  | 0 | 3,970 | 5,600 | 8,967 | 43,828 |
| **Remaining two-step algorithm tests** |  |  |  |  |  |
|  | 43,828 | 39,858 | 38,228 | 34,861 | 0 |
| **Antigen positivity (%) in combination antigen/antibody testing** |  |  |  |  |  |
|  | N/A | 3.55 | 3.19 | 3.64 | 0.93 |
| **Antibody positivity (%) in population undergoing two-step testing** |  |  |  |  |  |
|  | 4.23 | 3.51 | 3.35 | 2.34 | N/A |
| **Laboratory cost of combination antigen/antibody tests** |  |  |  |  |  |
| | \$0 | \$110,088 | \$155,288 | \$248,655 | \$1,215,350 |
| **Laboratory cost of HCV RNA tests reflexed from antibody-positive/antigen-negative tests** |  |  |  |  |  |
| | \$0 | \$15,303 | \$19,348 | \$35,013 | \$70,970 |
| **Laboratory cost of two-step algorithm testing** |  |  |  |  |  |
| | \$1,103,127 | \$989,231 | \$945,661 | \$845,033 | \$0 |
| **Total laboratory cost** |  |  |  |  |  |
| | \$1,103,127 | \$1,114,622 | \$1,120,296 | \$1,128,701 | \$1,286,320 |
| **Difference in laboratory costs compared to two-step algorithm** |  |  |  |  |  |
| | \$0 | \$11,494 | \$17,169 | \$25,573 | \$183,193 |
| **Laboratory cost per detected infection** |  |  |  |  |  |
| | \$2,473 | \$2,499 | \$2,512 | \$2,531 | \$2,884 |

**Table 4.** Cost analysis for switching to HCV antibody-to-antigen reflex testing based on implementation locations. The cost difference between using the traditional two-step antibody-to-RNA algorithm for HCV screening compared to HCV antibody-to-antigen reflex testing if the antibody test is positive. All values are based on estimated lab costs for HCV antibody testing, traditional HCV RNA testing, and Hepatitis B surface antigen testing which was utilized to estimate HCV antigen costs. All values are in US dollars. All values are based on 2024 values for reflex test orders, and costs are estimated for one year.

| Testing converted to antibody-to-antigen reflex tests | No additional testing | Public County Hospital ED | ED | Inpatient and ED | All Testing |
| --- | --- | --- | --- | --- | --- |
| **Number of antibody-to-antigen reflex tests** |  |  |  |  |  |
|  | 0 | 3,970 | 5,600 | 8,967 | 43,828 |
| **Remaining two-step algorithm tests** |  |  |  |  |  |
|  | 43,828 | 39,858 | 38,228 | 34,861 | 0 |
| **Antibody positivity (%) in antibody-to-antigen reflex tests** |  |  |  |  |  |
|  | N/A | 11.39 | 10.23 | 11.58 | 4.23 |
| **Estimated antigen positivity (%) for positive antibody results in population undergoing antibody-to-antigen reflex testing** |  |  |  |  |  |
|  | N/A | 31.16 | 31.30 | 31.41 | 22.13 |
| **Antibody positivity (%) in population undergoing two-step testing** |  |  |  |  |  |
|  | 4.23 | 3.51 | 3.35 | 2.34 | N/A |
| **Laboratory cost of antibody-to-antigen reflex tests** |  |  |  |  |  |
| | \$0 | \$93,942 | \$132,186 | \$212,271 | \$1,021,314 |
| **Laboratory cost of HCV RNA tests reflexed from antibody-positive/antigen-negative tests** |  |  |  |  |  |
| | \$0 | \$15,303 | \$19,348 | \$35,013 | \$70,970 |
| **Laboratory cost of two-step algorithm testing** |  |  |  |  |  |
| | \$1,103,127 | \$989,231 | \$945,661 | \$845,033 | \$0 |
| **Total laboratory cost** |  |  |  |  |  |
| | \$1,103,127 | \$1,098,475 | \$1,097,194 | \$1,092,317 | \$1,092,284 |
| **Difference in laboratory costs compared to two-step algorithm** |  |  |  |  |  |
| | \$0 | -\$4,652 | -\$5,933 | -\$10,811 | -\$10,844 |
| **Laboratory cost per detected infection** |  |  |  |  |  |
| | \$2,473 | \$2,463 | \$2,460 | \$2,449 | \$2,449 |

## Discussion

The recent FDA authorization of the first point-of-care HCV RNA test has generated significant enthusiasm among providers and policymakers as a potential catalyst for HCV elimination in the United States. However, questions remain about whether and how this test should be implemented across medical systems. To address this from a clinical laboratory perspective, we conducted a comprehensive analysis of recent HCV testing within our medical system. Overall, we observed a substantial increase in outpatient HCV antibody testing between 2019 and 2021, which was temporally associated with a local initiative to broadly screen for HCV as well as CDC’s 2020 recommendation of screening all adults for HCV in 2020, along with a corresponding decline in positivity rates. Despite this increase in antibody testing, viral load testing volumes have declined markedly since 2017, primarily in the outpatient setting, consistent with consolidation of earlier guideline changes that recommend viral load testing only at baseline and post-treatment cure assessment^29^.

With the availability of highly effective curative therapies and shifting risk factors for HCV, testing in the emergency department for HCV has gained increased attention nationwide^21,30^. In our medical system, emergency department testing rose in 2022 for both HCV antibody and viral load tests. The public county hospital emergency department accounted for the majority of these tests, reflecting a patient population with higher risk behaviors and less presentation to outpatient primary care. The antibody positivity rate in the emergency department was five times higher than the outpatient rate, indicating a significantly higher-risk population. Notably, in 2024, the positivity rate for reflex HCV RNA testing in the public county hospital emergency department was only 30.7%, underscoring the high proportion of previously treated individuals in this population, given spontaneous clearance rates are estimated to be between 15-40% for HCV infection^12^.

While testing is the first step toward curing HCV infection, patients must be linked to treatment to achieve improved clinical outcomes. A major limitation of the current testing algorithm is that HCV antibody tests reflexed to viral load confirmation do not yield results within the timeframe of an emergency department visit, as reflected by our median turnaround time of 84 hours from 2017 to 2024. No doubt this approach was originally designed for an outpatient-first testing model, rather than the current scenario where over one-third of HCV infections are diagnosed from the public county emergency department, as seen in our system. A recent study of HCV antibody screening in the emergency department found that fewer than 10% of newly diagnosed HCV infections were cured when only the antibody result was available during the emergency department visit, highlighting the potential importance of having the HCV RNA result during the visit^21^. To address turnaround time, we transitioned HCV viral load testing to the random access Hologic Panther platform, reducing median turnaround time to 45 hours, consistent with 1.6-4.0 day median turnaround time seen in laboratory-reflex based HCV testing ^31–33^. However, despite the instrument’s random-access capability, samples were still batched and run only three times weekly due to quality control costs associated with quantitative viral load testing. While this represents a significant improvement, the turnaround remains too long for a typical emergency department visit, indicating the need for alternative testing strategies.

In June 2024, the FDA authorized the first POC HCV RNA test, enabling results within the timeframe of a clinical visit. However, the costs associated with this testing approach remain high, particularly given strained hospital and Medicaid budgets. For example, we found that converting all HCV testing to this platform would increase laboratory costs by 260%, or approximately $2.87 million, across our three-hospital system. Restricting POC HCV RNA implementation to the public county hospital emergency department could substantially reduce turnaround time – other onsite Cepheid tests have median real-world turnaround times under three hours – for the 149 individuals diagnosed with HCV infection there. However, implementing this testing in just one hospital unit would increase annual laboratory costs by roughly a quarter of a million dollars, with likely increases as emergency department caseloads grow. These expenses may come at the cost of testing other patients amid constrained budgets.

While no HCV antigen tests are currently FDA-approved, antigen testing is used in other countries to confirm active HCV infection ^27,28,34^. We therefore modeled the same scenarios applied to POC HCV RNA testing but using either upfront combination HCV antigen/antibody testing or reflex antibody-to-antigen testing. These approaches resulted in much lower laboratory costs, and if on-site chemistry line testing is available, could potentially provide turnaround times equivalent to or better than POC HCV RNA testing. For example, implementing combination antigen/antibody testing in the public county hospital emergency department would add only $11,494 in costs – less than one-twentieth the additional cost of POC RNA testing – while reflex antibody-to-antigen testing was overall cost-saving from the laboratory perspective.

Our cost projections suggest that HCV antigen testing could be a cost-effective strategy for confirming active infection during an emergency department visit. While earlier smaller studies raised concerns about the analytical sensitivity of antigen tests^35,36^, our data indicate that antigen testing would miss only 2% of infections based on reflexed HCV viral loads. This aligns with previous reports showing 3%^37^, 5%^34^, and 6%^38^ of antigen tests resulting as negative despite detectable HCV viral loads. To avoid missing infections, we modeled an algorithm where all positive antibody but negative antigen tests are reflexed to HCV RNA testing, allowing eventual identification of these few cases, albeit with a longer turnaround time. While this reflex testing ensures no infections are overlooked due to lower antigen sensitivity, eliminating this reflex step could further reduce laboratory costs.

While our cost projections aimed to reflect realistic laboratory expenses, it has several limitations. We focused exclusively on laboratory costs and did include costs related to clinical care associated with this testing. We also did not factor in additional costs or savings on either healthcare or the broader societal level following cured HCV infections that do not progress to further complications. Our projections are based on 2024 testing data and does not include dynamic forecasting of future testing volumes. Because infectious diseases dynamics are nonlinear, successful or unsuccessful implementation of HCV test-and-treat strategies could result in different future infection trends, impacting future testing volumes and institutional costs^39^. Since no FDA-approved HCV antigen tests exist, we estimated costs using Hepatitis B surface antigen tests as a proxy, which may not be accurate. A key limitation is that our model considers positive test results rather than clinical outcomes. Linking patients to HCV treatment and cure remains a major challenge in emergency departments^21,40^, and must be addressed for the clinical benefits of testing to be realized. Despite these limitations, our findings align with a recent CDC study showing cost savings for an antibody-to-antigen reflex algorithm, while combined antigen/antibody testing or POC RNA testing were associated with higher costs^41^.

Another limitation of our study is that it focuses on a single medical system within one city. However, our findings are consistent with other published results, supporting their broader generalizability. For example, a study from Baltimore reported a 13.8% HCV antibody positivity rate in their emergency department, similar to our observed rate^30^. Additionally, our data show that 3.4% of emergency department tests resulted in confirmed HCV infections, which with rates reported in multi-city studies (1.9%)^21^ and a study from Baltimore (1.7%)^35^.

In summary, the recent availability of POC HCV RNA testing raises important questions about capacity, cost-effectiveness, and evolving HCV testing trends. Our data show that while emergency department testing is increasing within our system, it still represents only a small fraction of total HCV screening and can substantially increase laboratory costs. Thus, any testing strategy must balance the need for rapid results in the emergency department with the need for low-cost screening in outpatient populations. We demonstrate that an antibody-to-antigen reflex algorithm could result in laboratory cost savings while providing rapid results, assuming on-site testing is available. If an automated chemistry line with this capability is not available on-site, targeted POC RNA testing in high-positivity, rapid-turnaround settings offers an alternative approach. Furthermore, recent data from a large randomized control trial of emergency department HCV antibody screening – in which fewer than 10% of individuals achieved HCV cure over 18 months of follow-up – highlights the significant challenges in translating testing into successful health outcomes. Given the substantially higher costs of direct HCV RNA testing, enthusiasm for POC HCV RNA testing must be matched with rigorous studies evaluating its impact on HCV cure rates.

## Data Availability

Data available upon reasonable request

## Acknowledgements

We would like to thank Jonathan Reed, Dariia Vyshenska, Clarice Mauer, and Hee Jin Oh for helpful comments. This study received no external funding and was supported by the Department of Laboratory Medicine and Pathology. ALG reports contract testing to UW from Abbott, Cepheid, Novavax, Pfizer, Janssen and Hologic, research support from Gilead, outside of the described work.

## References

1. Hall, E. W. et al. Estimating hepatitis C prevalence in the United States, 2017-2020. Hepatol. Baltim. Md 81, 625–636 (2025).

2. Gnanapandithan, K. & Ghali, M. P. Self-awareness of hepatitis C infection in the United States: A cross-sectional study based on the National Health Nutrition and Examination Survey. PloS One 18, e0293315 (2023).

3. Hepatitis Surveillance in the United States, 2017 | CDC. CDC https://archive.cdc.gov/www_cdc_gov/hepatitis/statistics/2017surveillance/index.htm.

4. Bourlière, M. et al. Sofosbuvir, Velpatasvir, and Voxilaprevir for Previously Treated HCV Infection. N. Engl. J. Med. 376, 2134–2146 (2017).

5. Forns, X. et al. Glecaprevir plus pibrentasvir for chronic hepatitis C virus genotype 1, 2, 4, 5, or 6 infection in adults with compensated cirrhosis (EXPEDITION-1): a single-arm, open-label, multicentre phase 3 trial. Lancet Infect. Dis. 17, 1062–1068 (2017).

6. Puoti, M. et al. High SVR12 with 8-week and 12-week glecaprevir/pibrentasvir therapy: An integrated analysis of HCV genotype 1-6 patients without cirrhosis. J. Hepatol. 69, 293–300 (2018).

7. Hepatitis C Surveillance | 2023. CDC https://www.cdc.gov/hepatitis-surveillance-2023/hepatitis-c/index.html (2025).

8. Wester, C. et al. Hepatitis C Virus Clearance Cascade - United States, 2013-2022. MMWR Morb. Mortal. Wkly. Rep. 72, 716–720 (2023).

9. Cartwright, E. J., Patel, P., Kamili, S. & Wester, C. Updated Operational Guidance for Implementing CDC’s Recommendations on Testing for Hepatitis C Virus Infection. MMWR Morb. Mortal. Wkly. Rep. 72, 766–768 (2023).

10. CLFS Files. CMS https://www.cms.gov/medicare/payment/fee-schedules/clinical-laboratory-fee-schedule-clfs/files.

11. Aisyah, D. N., Shallcross, L., Hully, A. J., O’Brien, A. & Hayward, A. Assessing hepatitis C spontaneous clearance and understanding associated factors-A systematic review and meta-analysis. J. Viral Hepat. 25, 680–698 (2018).

12. Westbrook, R. H. & Dusheiko, G. Natural history of hepatitis C. J. Hepatol. 61, S58–68 (2014).

13. Bui, T. I. et al. Comparison of a dual antibody and antigen HCV immunoassay to standard of care algorithmic testing. J. Clin. Microbiol. 62, e0083224 (2024).

14. Mixson-Hayden, T. et al. Performance of ARCHITECT HCV core antigen test with specimens from US plasma donors and injecting drug users. J. Clin. Virol. Off. Publ. Pan Am. Soc. Clin. Virol. 66, 15–18 (2015).

15. New recommendation on hepatitis C virus testing and treatment for people at ongoing risk of infection Policy brief. WHO https://www.who.int/publications/i/item/9789240071872.

16. Viral Hepatitis Surveillance and Case Management - Appendices. CDC https://www.cdc.gov/hepatitis/statistics/surveillanceguidance/Appendices.htm (2025).

17. Liaw, W., Petterson, S., Rabin, D. L. & Bazemore, A. The impact of insurance and a usual source of care on emergency department use in the United States. Int. J. Fam. Med. 2014, 842847 (2014).

18. Cummings, E., Martinez, S. & Mourad, M. Primary care gap: factors associated with persistent lack of primary care after hospitalisation. BMJ Open Qual. 11, (2022).

19. Levine, D. M., Linder, J. A. & Landon, B. E. Characteristics of Americans With Primary Care and Changes Over Time, 2002-2015. JAMA Intern. Med. 180, 463–466 (2020).

20. FDA Permits Marketing of First Point-of-Care Hepatitis C RNA Test. FDA https://www.fda.gov/news-events/press-announcements/fda-permits-marketing-first-point-care-hepatitis-c-rna-test.

21. Haukoos, J. et al. Hepatitis C Screening in Emergency Departments: The DETECT Hep C Randomized Clinical Trial. JAMA (2025) doi:10.1001/jama.2025.10563.

22. Ivanova Reipold, E., Shilton, S., Donolato, M. & Fernandez Suarez, M. Molecular Point-of-Care Testing for Hepatitis C: Available Technologies, Pipeline, and Promising Future Directions. J. Infect. Dis. 229, S342–S349 (2024).

23. The UW Medicine Family. UW Medicine https://www.uwmedicine.org/about/the-uwmedicine-family.

24. Hepatitis C data dashboards - King County, Washington. King County Public Health https://kingcounty.gov/en/dept/dph/health-safety/disease-illness/hiv-sti-hcv/hepatitis-c/dashboard.

25. Schillie, S., Wester, C., Osborne, M., Wesolowski, L. & Ryerson, A. B. CDC Recommendations for Hepatitis C Screening Among Adults - United States, 2020. MMWR Recomm. Rep. Morb. Mortal. Wkly. Rep. Recomm. Rep. 69, 1–17 (2020).

26. European Association for the Study of the Liver. Electronic address, Clinical Practice Guidelines Panel: Chair:, EASL Governing Board representative:, & Panel members: EASL recommendations on treatment of hepatitis C: Final update of the series☆. J. Hepatol. 73, 1170–1218 (2020).

27. Mandel, E. et al. Province-to-province variability in hepatitis C testing, care, and treatment across Canada. Can. Liver J. 6, 234–248 (2023).

28. Handanagic, S. et al. Lessons Learned From Global Hepatitis C Elimination Programs. J. Infect. Dis. 229, S334–S341 (2024).

29. Bhattacharya, D., Aronsohn, A., Price, J. & Lo Re, V. Hepatitis C Guidance 2023 Update: AASLD-IDSA Recommendations for Testing, Managing, and Treating Hepatitis C Virus Infection. Clin. Infect. Dis. Off. Publ. Infect. Dis. Soc. Am. ciad319 (2023) doi:10.1093/cid/ciad319.

30. Hsieh, Y.-H. et al. Evaluation of the Centers for Disease Control and Prevention Recommendations for Hepatitis C Virus Testing in an Urban Emergency Department. Clin. Infect. Dis. Off. Publ. Infect. Dis. Soc. Am. 62, 1059–1065 (2016).

31. Tao, Y. et al. Laboratory-based and clinic-based hepatitis C virus viral load reflex testing following an initial positive HCV antibody test: a systematic review and meta-analysis. in Updated Recommendations on Treatment of Adolescents and Children with Chronic HCV Infection, and HCV Simplified Service Delivery and Diagnostics [Internet] (World Health Organization, 2022).

32. López-Martínez, R. et al. Significant Improvement in Diagnosis of Hepatitis C Virus Infection by a One-Step Strategy in a Central Laboratory: an Optimal Tool for Hepatitis C Elimination? J. Clin. Microbiol. 58, e01815–19 (2019).

33. Thompson, L. A., Fenton, J. & Charlton, C. L. HCV reflex testing: A single-sample, low-contamination method that improves the diagnostic efficiency of HCV testing among patients in Alberta, Canada. J. Assoc. Med. Microbiol. Infect. Dis. Can. 7, 97–107.

34. Pérez-García, A., Aguinaga, A., Navascués, A., Castilla, J. & Ezpeleta, C. Hepatitis C core antigen: Diagnosis and monitoring of patients infected with hepatitis C virus. Int. J. Infect. Dis. IJID Off. Publ. Int. Soc. Infect. Dis. 89, 131–136 (2019).

35. Prostko, J. et al. Performance evaluation of the Abbott Alinity Hepatitis C antigen next assay in a US urban emergency department population. J. Clin. Virol. Off. Publ. Pan Am. Soc. Clin. Virol. 175, 105743 (2024).

36. Wong, X. Z. et al. Hepatitis C core antigen testing to diagnose active hepatitis C infection among haemodialysis patients. BMC Nephrol. 21, 480 (2020).

37. Gunsolus, I. L. et al. Comparison of a hepatitis C core antigen assay to nucleic acid amplification testing for detection of hepatitis C viremia in a US population. Microbiol. Spectr. 12, e0097524 (2024).

38. van Tilborg, M. et al. HCV core antigen as an alternative to HCV RNA testing in the era of direct-acting antivirals: retrospective screening and diagnostic cohort studies. Lancet Gastroenterol. Hepatol. 3, 856–864 (2018).

39. Fraser, H. et al. Model projections on the impact of HCV treatment in the prevention of HCV transmission among people who inject drugs in Europe. J. Hepatol. 68, 402–411 (2018).

40. Hernandez-Con, P. et al. Hepatitis C Cascade of Care in the Direct-Acting Antivirals Era: A Meta-Analysis. Am. J. Prev. Med. 65, 1153–1162 (2023).

41. Hall, E. W. et al. Cost-Effectiveness Analysis of Testing Approaches for Diagnosis of Hepatitis C Among US Adults. Clin. Infect. Dis. Off. Publ. Infect. Dis. Soc. Am. ciaf166 (2025) doi:10.1093/cid/ciaf166.

